# Seroprevalence and preventive practices of dengue and chikungunya among school children in Bangkok: Gaps in prevention and vaccination strategies

**DOI:** 10.1101/2025.04.03.25325161

**Authors:** Thitiya Yakasaem, Thidarat Jupimai, Nattapong Jitrungruengnit, Napaporn Chantasrisawad, Ekasit Kowitdamrong, Padet Siriyasatien, Sunthorn Sunthornchart, Nattinee Isarankura Na Ayudaya, Paveena Angkhananukit, Pitsamai Ruansil, Kanchana Nakhapakorn, Eric Daudé, Alexandre Cebeillac, Richard Paul, Thanyawee Puthanakit, Watsamon Jantarabenjakul

**Affiliations:** Division of Pediatric Infectious Disease, Department of Pediatrics, Faculty of Medicine, Chulalongkorn University, Bangkok, Thailand; Center of Excellence for Pediatric Infectious Diseases and Vaccines Faculty of Medicine, Chulalongkorn University, Bangkok, Thailand; Thai Red Cross Emerging Infectious Disease Clinical Center, King Chulalongkorn Memorial Hospital, Bangkok, Thailand; Department of Microbiology, Faculty of Medicine, Chulalongkorn University, Bangkok, Thailand; Center of Excellence in Vector Biology and Vector Borne Disease, Department of Parasitology, Faculty of Medicine, Chulalongkorn University, Bangkok, Thailand; Pediatrics Department, Charoenkrung Pracharak Hospital, Bangkok, Thailand; Bangkok Metropolitan Administration, Bangkok, Thailand; Communication Disease Control Division, Health Department, Bangkok, Thailand; Public Health Service Center 24, Bangkok, Thailand; Department of Education, Bangkok, Thailand; Faculty of Environmental and Resource Studies, Mahidol, Bangkok, Thailand; Institut de recherche sur l’Asie du Sud-Est contemporaine IRASEC, CNRS, Bangkok, Thailand; Institut Pasteur, Université Paris Cité, CNRS UMR 2000, INRAE USC 1510, Ecology and Emergence of Arthropod-borne Pathogens Unit, Paris, France

**Keywords:** Dengue, Chikungunya, seroprevalence, prevention, KAP, Bangkok

## Abstract

**Background:** Dengue and chikungunya, both transmitted by *Aedes* mosquitoes, continue to pose significant public health concerns in Thailand, particularly during the rainy season. Despite ongoing vector control efforts, the incidence of infection remains high, with an increasing trend observed in chikungunya. This underscores the need for additional control measures, including vaccination, to reduce disease burden and morbidity. This study aims to assess the seroprevalence of dengue and chikungunya infections among children aged 10-15 years in Bangkok and to evaluate the knowledge, attitudes, and practices (KAP) related to mosquito-borne disease prevention.

**Methodology:** A cross-sectional descriptive study was conducted across 12 schools in Bangkok. Children aged 10 to 15 years were included. Seroprevalence was determined using rapid diagnostic tests (Abbott DENGUE IgG/IgM and Citest Chikungunya IgG/IgM) based on the immunochromatography technique, using fingertip blood samples. Parents completed KAP questionnaires, including factors influencing vaccination decisions.

**Principal findings:** From June to August 2024, 937 participants were enrolled, with a mean (SD) age of 11 (1.6) years; 67% were aged 10–12 years, and 33% were aged 13–15 years. The seroprevalence of dengue was 28.1% (95% CI 25.2-31.0), while that of chikungunya was 6.3% (95% CI 4.7-7.9). KAP assessments revealed a high level of awareness regarding symptoms and transmission; however, notable deficiencies in preventive behaviors were identified. Only 14.8% of respondents reported consistent use of mosquito repellent, and 17.5% routinely inspected and removed mosquito larvae from their homes.

**Conclusion:** The substantial seroprevalence of dengue and the emerging trend of chikungunya among children in Bangkok highlights the urgent need to enhance community education and strengthen vector control interventions. Expanding dengue vaccination coverage and raising awareness about chikungunya prevention, including consideration for future vaccine implementation, are essential to mitigating future outbreaks and reducing the disease burden.

**Author summary:**

- Mosquito-borne diseases, including dengue and chikungunya, are significant health problems in Southeast Asia. In our study, we evaluate the seroprevalence of these diseases using rapid blood tests in a school-based setting among children in Bangkok. The seroprevalence data highlight the risk of exposure, particularly among children, and can guide preventive practices, including vaccination. Despite the widespread recognition of these diseases, preventive practices remain limited. Therefore, implementing effective preventive measures and vaccination strategies could significantly reduce the severity of infections and improve public health outcomes.

## Introduction

Mosquito-borne diseases, particularly dengue and chikungunya, remain significant public health concerns in Southeast Asia. As of October 2024, the reported incidence of dengue was 156 cases per 100,000 population, with Bangkok recording a morbidity rate of 115 per 100,000. ^(1)^ In 2023, the highest infection rates were observed in children and adolescents.^(2)^ A systematic review reported seroprevalence among Thai children aged 2-17 years, assessed by the plaque reduction neutralization test (PRNT), reaching up to 80% in certain regions. ^(3)^ The seroprevalence increases with age; among 2–4-year-olds, reports indicate 48%, 5-8 years report 61.9%, 9-12 years report 79.5 and 13-16 year report 84.2%. ^(4)^ Chikungunya, while less prevalent than dengue, is more commonly reported in adolescents and adults, with symptoms including rash, severe joint pain, and potential neurological complications in children.^(5)^ A systematic review study covering the African, American, and Southeast Asian regions found a seroprevalence of 7% in children under five years old, consistent with findings from a 2014-2015 study [6,7]. Chikungunya outbreaks have been more frequent in Thailand in southern regions. A study in 2017 reported a seropositivity rate of Chikungunya IgG antibodies by commercial enzyme-linked immunosorbent assays (ELISA) of 3% among individuals aged 10–19 in central Thailand compared to 20% in the South.^(8)^ However, data on Chikungunya seroprevalence in Thai children remains limited.

Given that *Aedes* mosquitoes transmit both diseases, their spread is influenced by social and environmental factors. While prevention measures such as vector control and personal protection are widely promoted, adherence to these practices varies. Effective disease prevention relies on public knowledge, attitudes, and practices (KAP). A cross-sectional study in the South of Thailand found that children previously infected with dengue exhibited better preventive practices, and over 70% of children cited teachers as their primary source of dengue information.^(9)^ Parental engagement in vector control efforts may further strengthen prevention efforts, and access to vaccination and other interventions also impacts disease control efforts. Despite ongoing vector control measures and the introduction of two dengue vaccines, Dengvaxia^®^ in 2017 and Qdenga^®^ in 2023, dengue incidence continues to rise, particularly after the COVID-19 pandemic. The WHO recommends dengue vaccination in high-prevalence areas. ^(10)^ Currently, the Chikungunya vaccine is available when there is an outbreak or when there is evidence of viral transmission within the past five years, especially in older adults with co-morbidities. ^(11)^

This study aims to determine the seroprevalence of dengue and chikungunya among children in Bangkok and assess parental KAP regarding disease prevention.

## Materials and methods

### Study design and inclusion/exclusion criteria

A cross-sectional observational study was conducted in 12 schools in Bangkok. The study included children aged 10-15. Children with medical conditions that could interfere with blood sampling on the study day were excluded.

### Ethical statement

Written assent was obtained from participants whose parents provided informed consent for blood sampling. Parents who consented to participate were included in the questionnaire-based survey. This study was part of the Geomosquito study and was approved by the Institut Pasteur Institutional Review Board (N°2023-136), the Chulalongkorn Institutional Review Board (IRB No. 0648/66), and the Bangkok Metropolitan Administration (BMA) Human Research Ethics Committee (BMAHREC No. E005hc/67). It was registered with the Thai Clinical Trials under the number TCTR20240404002.

### Study procedures

The Bangkok metropolitan area was divided into five regions: Northern, Southern, Eastern, Western, and Central. Schools were selected from each region after initial contact and agreement to participate. The study was conducted on school premises, where parents provided consent for their children’s participation and completed questionnaires to screen history and assess their knowledge, attitudes, practices, and factors related to dengue and chikungunya vaccination. Students received study details, assent, and parent consent documents, and they participated in an educational session about dengue diseases and prevention. Blood samples were collected via fingertip capillary blood sampling for dengue and chikungunya antibody testing.

### Dengue and Chikungunya screening test

Rapid diagnostic tests were used to detect antibodies. The Abbott DENGUE IgG/IgM (Abbott Laboratories, USA) was used for dengue, while the Citest IgG/IgM (CITEST diagnostic inc., Canada) was used for chikungunya. Both tests utilize immunochromatography, requiring only 10 microlites for IgG and IgM detection. The reported sensitivity and specificity for the dengue test are 94.2% and 96.4%, while for the chikungunya test, they are 90.3% and 99.9% for diagnosing acute illness, respectively.^(12–13)^ Blood samples were collected using a capillary tube and transferred to the test kit with buffer, and results were available in 15-20 minutes. Two trained technicians independently interpreted the results, with a third confirmation if the results were discordant.

### Questionnaire development

A structured questionnaires was used to collected demographic data and assess parents’ knowledge, attitudes, practices, and factors influencing vaccination uptake for dengue and chikungunya. The questionnaire consisted of 28 questions and was designed to take 3-5 minutes to complete. The questionnaire was reviewed by physicians to ensure content validity, and its reliability was assessed using Cronbach’s alpha.

### Sample size calculation

The sample size was determined based on expected seroprevalence rates of dengue and chikungunya from previous studies. For dengue, seroprevalence ranges from 18% to 84%, requiring a sample size of approximately 207 to 227 participants. For Chikungunya, a prevalence of 3% suggested a minimum sample size of 45 participants. To ensure adequate statistical power, a larger sample size was chosen. Assuming a dengue infection seroprevalence of 55%, with a margin of error of 5% and a 5% alpha error, the calculated sample size was 380. With that sample size, if chikungunya seroprevalence is 3%, the sample size should be 1000 and the study would have over 99% power to detect it. However, this study was a substudy of a geomosquito study, which included at least 840 partipants; we enrolled according to the main study.

### Data management and analysis

For each participant, data on age, sex and history of infection were collected. The seroprevalence, defined by positive IgG, was calculated overall and by region, with 95% confidence intervals (CI). The history of dengue vaccination was reported as a percentage. Differences between age groups (10-12 and 13-15 years) and regions (five regions) were analyzed using Chi-square tests and one-way ANOVA. Vaccine knowledge and willingness to vaccinate were presented as percentages. Multivariate regression analysis was conducted using generalized linear models to identify factors associated with vaccine uptake. Data management utilized REDCap version 13.7.25 and was analyzed by SPSS version 29.0.0.0 (241).

## Result

### Demographic data

From June to August 2024, a total of 937 students were enrolled, with a mean age of 11 years (SD: 1.6). The demographic characteristics are summarized in Table 1. Schools were recruited from all regions of Bangkok, including two from the Central, one from the East, one from the North, two from the West, and six from the South.

**Table 1.**
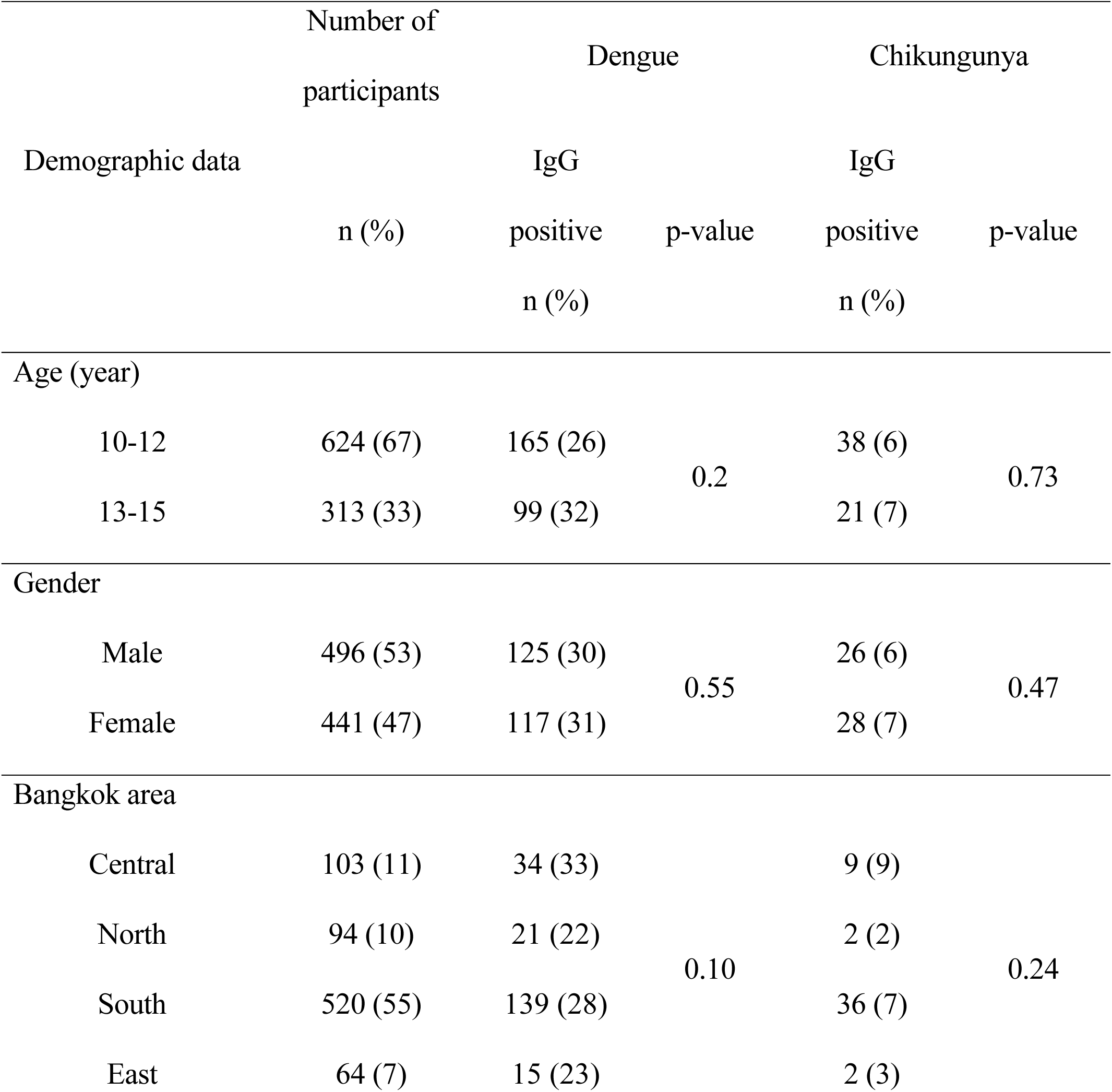
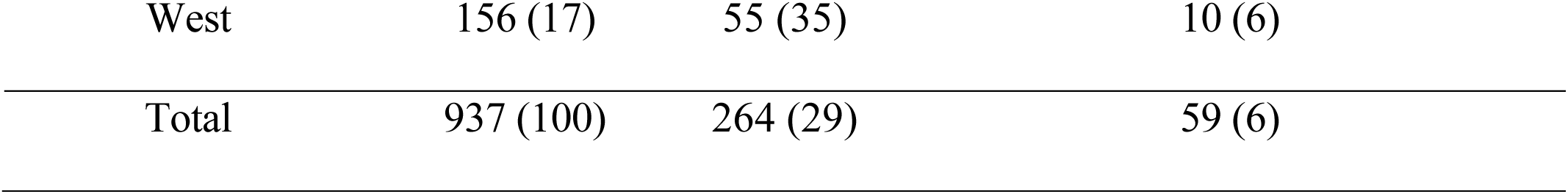
Demographic data and seroprevalence of Dengue and Chikungunya in children aged 10-15 years across Bangkok (N = 937).

The questionnaire study included responses from 889 participating parents, representing a 95% response rate. The mean age of respondents was 42 years (SD: 8.8). Most parents (65%) had resided in their current neighborhood for over five years. Regarding housing type, 34% of participants lived in detached houses, 25% in condominiums or apartments, 19% in townhouses, and 17% in other accommodations such as school dormitories or camps. Parental education levels were distributed as follows: 16% had completed primary school, 35% secondary school, and 23% held a bachelor’s degree or higher. Additionally, 4% had a vocational certificate (a specific practical training program equivalent to a post-secondary qualification), while 22% did not provide information on educational attainment.

### Prevalence of dengue and chikungunya infection

The overall seroprevalence, determined by positive IgG results, was 28.1% (95% CI: 25.2–31.0) for dengue and 6.3% (95% CI: 4.7–7.9) for chikungunya (Table 1 and Fig 1). Dengue IgM positivity was observed in 7% of participants, while chikungunya IgM was positive in 0.2%. None of the IgM-positive participants reported fever at the time of blood collection. Dengue seroprevalence exhibited a tendency to increase with age, whereas no such trend was observed for Chikungunya seroprevalence. Regional variations in seroprevalence were observed, though these differences were not statistically significant. The highest dengue seroprevalence was recorded in the western region of Bangkok (35.3%, 95% CI 32.2-38.4), while the lowest was in the northern region (22.1%, 95% CI 19.4-24.8). Chikungunya seroprevalence was highest in Central Bangkok (8.7%, 95% CI 6.9-10.5) and the lowest in the northern region (2.1%, 95% CI 1.2-3.0).

**Fig 1.**
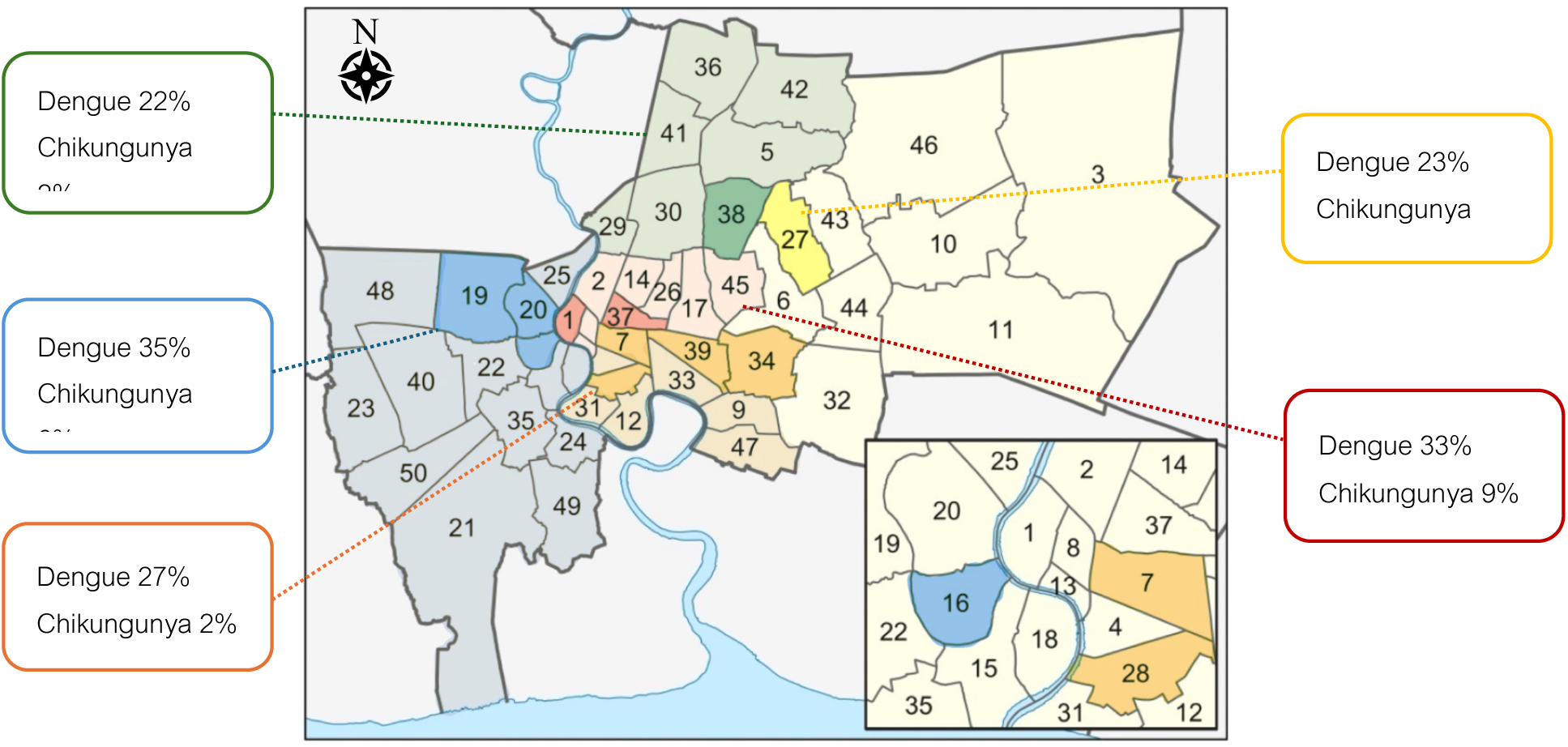
Seroprevalence of Dengue and Chikungunya by geographic area among Bangkok youth aged 10-15 years (N = 937) (Each color represents a zone in Bangkok: red for central, green for north, orange for south, yellow for east, and blue for west. Darker shades indicate the study areas where seroprevalence data was collected.)

Thirty-seven participants (3.9%) reported a previous dengue infection confirmed by physicians or blood testing, while four participants (0.4%) reported a prior chikungunya infection. Among those with a documented history of dengue infection, 46% tested positive for dengue IgG, compared to 27% of those without a reported history of infection. Notably, all four participants with a prior history of Chikungunya infection tested positive for Chikungunya IgG.

### Knowledge, attitude, and practice regarding dengue and chikungunya

KAP assessments revealed that most parents demonstrated a high level of awareness regarding dengue and chikungunya transmission and symptoms, with 71-94% correctly responding to dengue-related questions and 69-81% correctly answering chikungunya-related items (Table 2). However, only 58% were aware of the availability of dengue vaccines, and just 56 participants (6.6%) reported their child had received it.

**Table 2.** Questionnaires on knowledge of dengue and chikungunya infection (N = 889*)

| Questionnaires on knowledge of dengue and chikungunya infection | Yes (%) | No (%) |
| --- | --- | --- |
| 1. *Aedes* mosquitoes are carriers of dengue fever | 885 (94) | 4 (1) |
| 2. The rainy season is the period with the highest outbreak of dengue fever | 876 (93) | 13 (2) |
| 3. The mosquitoes that cause dengue fever are active during daytime | 667 (71) | 222 (24) |
| 4. Dengue fever can cause a high fever lasting 2-7 days, along with symptoms like abdominal pain and nausea | 830 (89) | 59 (6) |
| 5. Dengue fever can be transmitted through contact with an infected person | 193 (21) | 692 (74) |
| 6. Currently, there is no vaccine to prevent dengue fever | 344 (37) | 544 (58) |
| 7. Fever, joint pain, and red eyes are symptoms of chikungunya | 762 (81) | 126 (13) |
| 8. In Chikungunya, joint pain can last for months | 651 (69) | 236 (25) |

\*The number of parent participants varied depending on the completeness of their questionnaires.
(The correct answers highlighted in green)

Regarding disease perception, 95% of parents recognized the potential severity of dengue, while 91% considered it a significant public health concern. However, 15-20% remained uncertain about the severity and health impact of chikungunya infection (Fig 2a).

**Fig 2:**
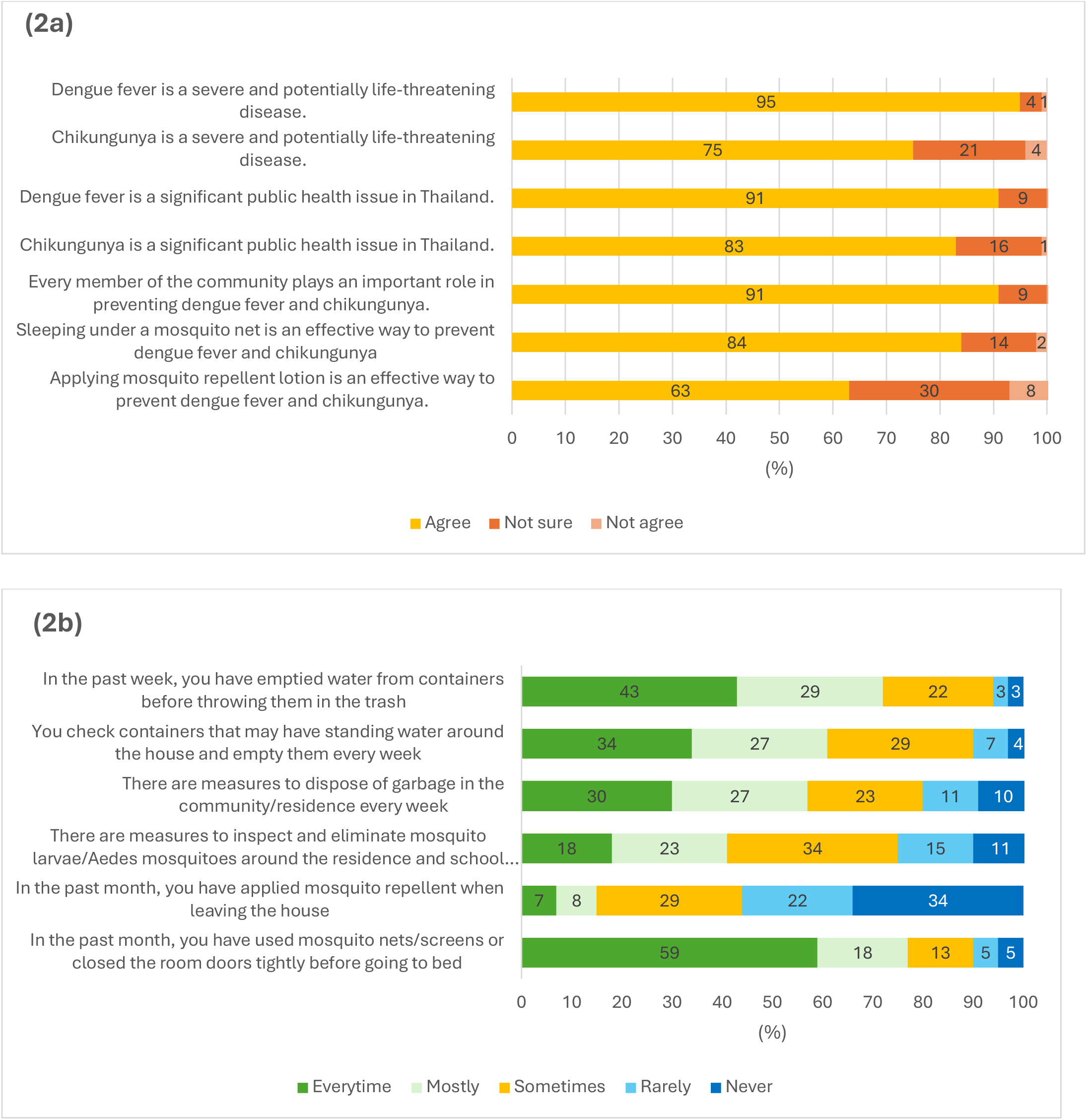
Questionnaire on Attitudes(2a) and Practices(2b) about Dengue and Chikungunya infection and prevention.

With respect to attitudes toward preventive measures, 63% of parents believed mosquito repellent was an effective prevention method, and 91% considered sleeping under mosquito nets as effective. The majority also agreed that community involvement was essential for disease control (Fig 2a). However, actual preventive practices were suboptimal; only one-third regularly checked for standing water, and 10% reported no preventive measures being taken in their residential areas. Additionally, 44% of parents used mosquito repellent before going outside, whereas 34% reported never using it (Fig 2b). There was no statistically significant difference in seroprevalence between individuals who regularly implemented preventive measures and those who did not (31%, 95% CI 30.4-30.9 vs 28%, 95% CI 28.0-28.3; p=0.10).

Most parents reported interest in vaccination, with 96% of parents expressing willingness to receive the dengue vaccine, and 94% were interested in a chikungunya vaccine. All parents of participants with a history of dengue or chikungunya infection were willing to receive the respective vaccines. Among those without a prior history of infection, 96% expressed interest in the dengue vaccination, while 94% indicated willingness to receive a chikungunya vaccination. The primary factors influencing vaccine uptake were safety (65-68%) and efficacy (58-61%). Cost was the least influential consideration, with only 31-34% citing it as a significant concern (Fig 3).

**Fig 3.**
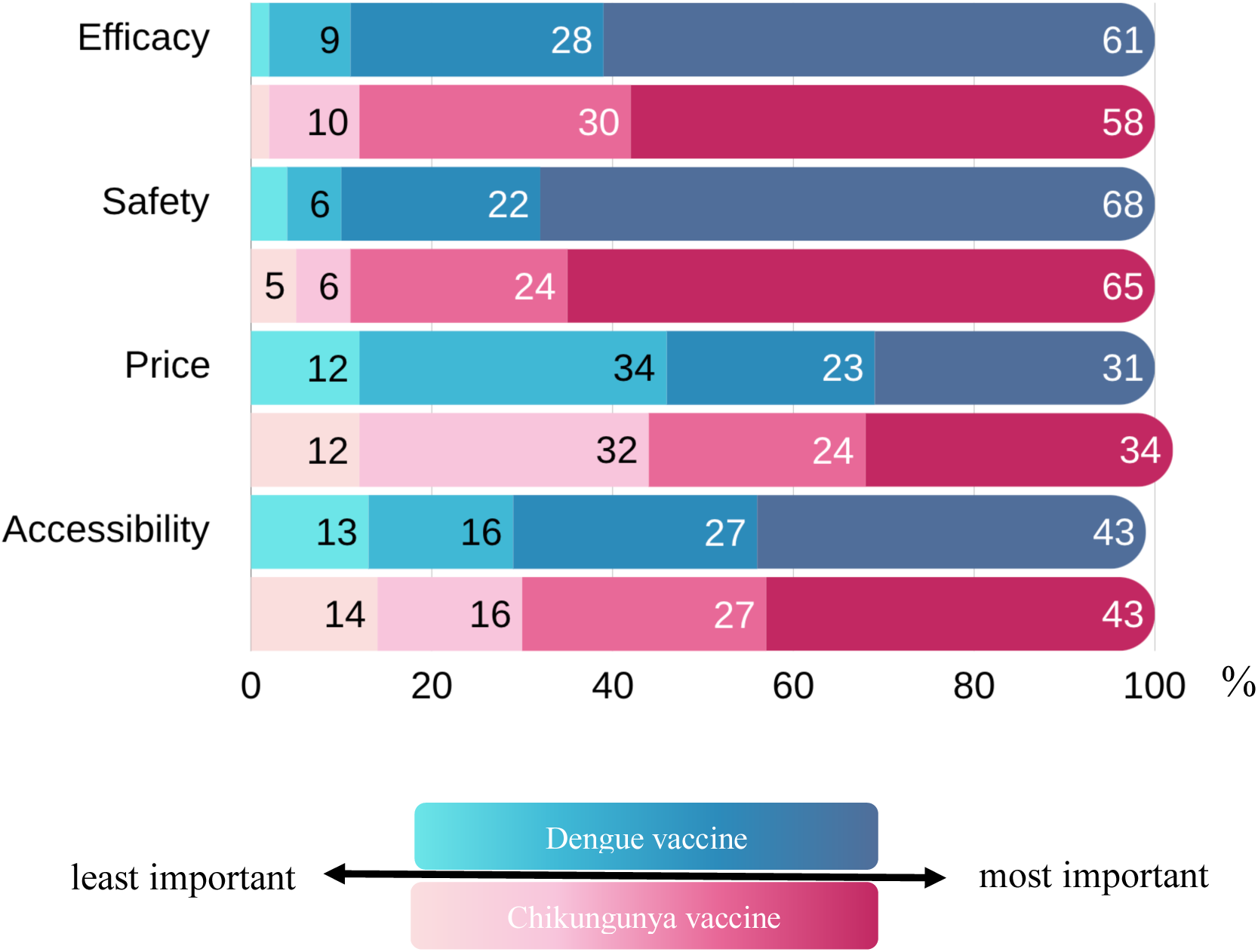
Factors that influence the reception of dengue and chikungunya vaccines.

## Discussion

The study provides significant insights into the seroprevalence of dengue and chikungunya among children aged 10-15 years in Bangkok. The observed dengue seroprevalence of 28.1% and chikungunya seroprevalence of 6.3%, as determined by rapid diagnostic tests, underscore the continued transmission of these arbovial infections in Bangkok. Notably, only 4% of participants reported a prior dengue infection, despite the higher measured seroprevalence, suggesting a substantial proportion of asymptomatic or mild cases among children. This finding raises concerns about the risk of severe dengue infections upon secondary infection, emphasizing the need for enhanced surveillance and preventive strategies. Furthermore, the report dengue seroprevalence in this study is lower than that documented in a 2023 study, which estimated a seroprevalence of 55.4% (95%CI 54.3-56.4) in the 10-14 age group using ELISA.^(14)^ This discrepancy may arise from differences in diagnostic methods, as ELISA exhibits higher sensitivity in detecting past infections compared to the rapid diagnostic test, which primarily identifies acute or recent infections. Among participants with a self-reported history of dengue infection, 46% tested positive for dengue IgG, suggesting that the true seroprevalence may be nearly double, aligning more closely with prior studies.

The chikungunya seroprevalence was higher than that reported in a study conducted in 2017, which reported a seropositivity of 3% among individuals aged 10–19 using ELISA. ^(8)^ The rising chikungunya seroprevalence in Bangkok may be attributed to the nationwide outbreak in 2019. This outbreak, particularly prominent in Bangkok, was associated with mutations in chikungunya virus strains, such as the E1-A226V mutation, which enhances viral adaptation to *Aedes aegypti* mosquitoes. Previous researches suggest that such genetic adaptations contribute to increased transmission and prolonged outbreaks. ^(15–16)^ Given these trends, continuous surveillance and research are essential to understanding the epidemiological shifts in chikungunya and informing targeted intervention strategies.

While regional variations in seroprevalence were observed, these differences were not statistically significant, likely due to regional heterogeneity in infection dynamics and sample size limitation. Variability in dengue prevalence across Bangkok aligns with prior findings, where the highest prevalence was reported in the southern region of Bangkok in 2023. ^(17)^ Differences in environmental conditions, population density, and mosquito control measures likely contribute to these disparities, warranting further investigation through spatial epidemiological studies in the Geomosquito study.

The KAP assessment demonstrated a high level of awareness regarding dengue and chikungunya among parents, with most recognizing these diseases as significant public health concerns. Knowledge levels were generally consistent across different regions and educational backgrounds, a finding that contrasts with a study in Singapore, where KAP scores varied by education levels. ^(18)^ Despite high awareness, gaps in preventive practices were evident. Barriers such as limited access to mosquito repellent, a lack of community-wide control initiatives, and socioeconomic constraints may hinder the consistent adoption of preventive behaviors. Additionally, urbanized living environments may make it more difficult for residents to eliminate standing water effectively. Only half of the parents regularly used mosquito repellent, and approximately one-third reported routinely checking for standing water – both crucial practices for mosquito control. The inconsistent use of mosquito repellent may stem from a perceived low risk or forgetfulness.

Despite relatively high awareness of dengue vaccines, uptake remained low, with only 6.6% of children vaccinated. This low coverage may be attributed to concerns regarding vaccine safety and efficacy and barriers related to accessibility and cost. Although chikungunya awareness was comparatively lower, more than 90% of parents expressed interest in chikungunya vaccination. Current vaccine recommendations primarily target travelers to endemic areas or populations in endemic regions.^(11)^ However, given the rising chikungunya burden in Thailand, post-outbreak seroprevalence studies and consideration of target vaccination strategies are warranted.

Notably, this study found no significant difference in seroprevalence between individuals who adhered to preventive measures and those who did not. This finding underscores the need for additional strategies, such as vaccination, to reduce the disease burden effectively. Although dengue vaccination is effective against severe dengue, the vaccine remains optional in Thailand. Safety and efficacy were the primary concerns among parents, outweighing cost considerations. Given the endemic nature of dengue in Thailand and the exclusion of dengue vaccines from the national immunization program, public health initiatives should focus on expanding vaccine accessibility and targeting high-risk populations to mitigate severe disease outcomes.

The strengths of this study include its large sample size, the specific focus on Bangkok, and an age-specific analysis of children aged 10-15 years, enhancing the reliability and applicability of the findings. In addition to assessing KAP, the study provides insights into vaccination acceptance, which is crucial for informing future vaccination strategies. Furthermore, the study actively promotes vector control awareness by providing education sessions. However, several limitations should be acknowledged. The study population was not evenly distributed across Bangkok, which may limit the generalizability of the findings to the entire city. Additionally, the use of rapid diagnostic tests, which have lower sensitivity compared to ELISA or PRNT, may have underestimated seroprevalence, particularly for past infections where antibody levels decline over time. The selection of rapid diagnostic tests was based on logistical feasibility within a school-base setting, as they require fingertip samples. Future studies employing ELISA or PRNT could provide more precise estimates of the seroprevalence for these mosquito-borne diseases.

## Conclusion

Dengue and chikungunya infections remain significant public health concerns in Thailand, with annual outbreaks and a rising chikungunya trend. Despite high awareness of these diseases, preventive practices are not consistently adopted, likely due to urban living constraints and socio-economic factors. The low uptake of dengue vaccination, despite its demonstrated efficacy, underscores the need for stronger public health initiatives to promote vaccine acceptance and accessibility.

## Data Availability

All relevant data are within the manuscript and its Supporting Information files.

## Acknowledgement

Division of pediatric infectious disease, Department of pediatrics, faculty of medicine, Chulalongkorn university. Center of excellence for pediatric infectious diseases and vaccines, Chulalongkorn university. Affiliated researchers at the faculty of medicine, Chulalongkorn university. Faculty of environmental and resource studies, Mahidol university. Institut de recherche sur l’Asie du Sud-Est contemporaine IRASEC, CNRS, Bangkok. Institut Pasteur, Paris, France. Cassandre von Platen, Institut Pasteur Pole for coordination of clinical research.

